# Protocol for the development of a core outcome set for the evaluation of shared decision-making interventions in healthcare

**DOI:** 10.64898/2025.11.28.25341200

**Authors:** Christin Hoffmann, Joanne Butterworth, Florian Naye, Maureen Smith, Taona Nyamapfene, Samuel Lawday, Kerry Avery, Hilary Bekker, Paulina Bravo, Simon Décary, Adrian Edwards, Glyn Elwyn, Ellen Engelhardt, Juan Franco, Mirjam Garvelink, Anik Giguere, Martin Härter, Tammy Hoffmann, Simone Kienlin, Kirsten McCaffery, Janneke Noordman, Karina Olling, Lilisbeth Perestelo-Perez, Arwen Pieterse, Fülöp Scheibler, Karen Sepucha, Dawn Stacey, Dirk Ubbink, Katherene Valentine, Robert Volk, Felix Wehking, Sang-Ho Yoo, Helen Bulbeck, Amy Cole, Maarten de Wit, Jeanette Finderup, Katie Geary, Christine Gunn, Wen-Hsuan Hou, Ashley Housten, Min Ji Kim, Norma Martí, Lissa Pacheco-Brousseau, Lisbeth Snede, Yue Tian Cindy, Karine Toupin-April, Angus McNair

**Affiliations:** University of Bristol, UK; University of Exeter, UK; Université Laval, Canada; Patient Partner, Ottawa, Canada; University Hospitals Plymouth NHS Trust, UK; University of Leeds, UK; Fundación Arturo López Pérez and Universidad Finis, Santiago, Chile; Université de Sherbrooke, Québec, Canada; Cardiff University, UK; Dartmouth College, USA; Netherlands Cancer Institute, The Netherlands; Heinrich-Heine-University Düsseldorf, Düsseldorf, Germany; St Antonius Hospital, Nieuwegein, The Netherlands; Laval University, Canada; University Hospital Hamburg-Eppendorf, Germany; Bond University, Australia; The South-Eastern Norway Regional Health Authority, Hamar, Norway; University of Sydney, Australia; Nivel (Netherlands Institute for Health Services Research), The Netherlands; Center for Shared Decision Making Vejle-Lillebaelt University Hospital of Southern Denmark; Canary Islands Health Service: Servicio Canario de Salud, Tenerife, Spain; Leiden University Medical Center, The Netherlands; University Hospital Cologne, University Hospital Schleswig-Holstein, Germany; Harvard University, USA; University of Ottawa, Canada; Amsterdam University Medical Center, The Netherlands; The University of Texas MD Anderson Cancer Center, USA; University Hospital Jena, Germany; Hanyang University, South Korea; Patient and public representative, University of Exeter, UK; The University of North Carolina at Chapel Hill, USA; Patient and public representative, The Netherlands; Department of Clinical Medicine, Aarhus University, Aarhus, Denmark; University of Warwick Medical School, UK; Taipei Medical University, Taiwan; Washington University School of Medicine, USA; Patient and public representative, North Carolina, USA; President, PiCC United (Patient Involvement & Collaboration Community United), Denmark; The Chinese University of Hong Kong, China

## Abstract

**Introduction:** Shared decision-making (SDM) is a process whereby patients are supported to reach decisions about their healthcare in collaboration with healthcare professionals. International policy and clinical guidelines highlight the ethical imperative of SDM and recommend SDM for many healthcare decisions and contexts. However, despite decades of SDM research, the impact of implementing SDM interventions within health care remains uncertain. High-quality health technology assessment (HTA) requires an understanding of how interventions to facilitate the adoption and implementation of SDM (e.g., through the use of patient decision aids, decision coaching, question prompt lists, training and feedback, or service changes) impact clinical and health service outcomes. Yet, synthesis of the existing literature is hindered by substantial heterogeneity in the evaluation of interventions to facilitate SDM. A core outcome set (COS) is an agreed standardised set of outcomes that should be measured and reported in all effectiveness studies. There is a COS for SDM in the context of rheumatology (rheuCOS-SDM), designed for use in research studies (e.g., clinical trials or observational studies) evaluating the impact of SDM interventions on clinical outcomes for patients. It is unclear, however, whether the outcome domains identified within a rheumatology context are relevant, comprehensive, or comprehensible when applied to a variety of SDM interventions tailored to and interacting with a range of patient populations, healthcare settings and contexts.

The aim of this study is to develop a generic COS for evaluating the impact of SDM interventions on various outcomes. Outcomes for consideration may include assessments of the behaviours and experiences of patients, important others (e.g., carers or relatives) and health professionals, the dynamics within patient-professional interactions, health outcomes for individuals and for the wider population, and the cost-effectiveness of care. The broad scope of the COS will ensure its applicability and utility within diverse healthcare contexts and enable the synthesis of evidence to draw clear conclusions about the impact of SDM interventions, to influence healthcare policy. We define this new, comprehensive COS as the COS-SDM.

**Methods and Analysis:** Through engagement with key interest holders (including patients and members of the public, clinicians and academics), we agreed on the scope of the COS and to adhere to the Core Outcome Measures in Effectiveness Trials (COMET) handbook and Core Outcome Set-STAndards for Development (COS-STAD) guidelines. This will involve production of a ‘long’ (comprehensive) list of candidate outcome domains (using evidence synthesis, a COS developed in the context of Rheumatology, and qualitative interviews with interest holders internationally), prioritisation of a ‘short’ (refined) list of core outcome domains (utilising a sequential two-round international online Delphi), and reaching consensus on the final outcome set (through international meetings, applying modified nominal group techniques and predefined criteria for agreement).

**Ethics and Dissemination:** Research ethics approval has been granted in the UK (University of Bristol Faculty Ethics Committee, ref: 7741; University of Exeter Faculty Ethics Committee, ref: 8207624).

The final COS will be disseminated by presentation at international conferences and publication in a peer-reviewed journal. Further dissemination is planned through our patient/public advisory group, professional networks, and executive group channels, to publicise the COS to patient groups, funders, journal editors, international regulatory bodies and HTA boards.

**Registration:** This project has been registered in the COMET database (www.comet-initiative.org/Studies/Details/3586).

## Introduction

### Background and context

Shared decision-making (SDM) is a process where patients are supported to reach decisions about their healthcare in collaboration with healthcare professionals (1). SDM involves patients, professionals, and others working together to choose care based on scientific evidence, clinical expertise, and personal preferences and values. International policy (2-4), professional and regulatory guidelines (5, 6) recommend SDM for a wide range of healthcare decisions in a variety of clinical contexts. SDM is recognised as a patient’s right, has an ethical imperative and inherent social value (2, 7).

Individuals often report that the process of SDM is a meaningful experience (8). High-quality health technology assessment (HTA), however, requires an understanding of how interventions to facilitate the adoption and implementation of SDM impact clinical and health service outcomes beyond process and patient experience (9). SDM interventions are often delivered through behaviour change (10), targeting one or more individuals who can be viewed as acting at various levels of a patient-centred healthcare system (i.e., interventions may work by changing the behaviours of patients themselves, or those of healthcare professionals, service managers, commissioners, or policy makers). There are many types of SDM intervention that influence the behaviours of relevant individuals through a variety of practical applications (e.g., through the implementation of patient decision aids, decision coaching, question prompt lists, training and feedback, or service changes). Some complex SDM interventions may have multiple, interacting components, targeting more than one individual acting at multiple levels (11).

Potential outcomes of these different SDM interventions range from those that assess SDM process (including patient and professional experiences), through those that assess immediate effect (e.g., aspects of patient health status), to those that relate to the longer term (e.g., patient self-management). Outcomes reported in the existing literature include patient knowledge of available options and informed preferences, realistic patient expectations about their care, improved decision self-efficacy, less decisional conflict, improved collaborative deliberation between patient and professional, better alignment between a chosen management option and a patient’s values in the context of their daily lives, reduced risk and impact of harm to patients, improved patient self-management, increased use of treatments with greater benefit-to-harm ratios, a reduced preference for the most intensive management plans, and appropriate health care utilization (12-28).

One outcome of SDM may influence another, i.e., benefits to patient and/or healthcare professional may in turn lead to more effective and cost-effective care. Evidence is emerging to enable the testing of mechanisms underlying the components of SDM interventions, to enable the mapping of non-linear, interacting pathways from process through to outcomes (29). Some evidence suggests that there might be longer-term impacts on health services. SDM interventions may reduce unwarranted practice variation and costs by reducing the rate that patients opt for unnecessary, expensive treatments (30). A systematic review identified 15 studies (11 randomized controlled trials (RCTs); 10 low risk of bias) that evaluated the impact of decision aids on these outcomes (31). Three demonstrated statistically significant reductions in rates of prostatectomy, hip/knee replacement, and hysterectomy (31). Initiatives, such as those from the UK Academy of Medical Royal Colleges and the former UK National Health Service for England’s Evidence-based Interventions programme, promote SDM to reduce patient harm and to improve use of resources (32). High-quality SDM may also reduce litigation, the associated emotional burden on patient and professional, and the resultant financial costs to the health care system (33, 34).

#### Rationale

Despite decades of research on SDM, the effect of SDM interventions on outcomes relating to patients, professionals, organisations, health systems, and populations, remains largely uncertain. Evidence synthesis/meta-analysis is hindered by a lack of (i) differentiation between SDM process measures and outcome measures, (ii) consensus on the priority outcomes of SDM (35), (iii) transparency when selecting these outcomes, and (iv) interpretability of assessments. This results in methodological heterogeneity, outcome reporting bias and research waste.

Efforts have been made to address these problems through the development of a core outcome set (COS) for use when evaluating SDM interventions in rheumatology (hereafter defined as the rheuCOS-SDM) (36). A COS is an agreed standardised set of outcomes that should be measured and reported, as a minimum, in all comparative effectiveness studies in specific areas of health or health care (37). The rheuCOS-SDM was co-developed with significant input from patients, clinicians and SDM experts in the field of rheumatology, using accepted methods to gain consensus on the COS within a rheumatology setting. It does, however, have some limitations. Initial development lacked clear conceptualisation to differentiate between SDM process and outcome assessment, although this was later clarified. The generation of a ‘long’ (comprehensive) list of outcome domains predated relevant, comprehensive systematic reviews (38, 39) that may have subsequently identified outcome domains to be shortlisted for inclusion in the consensus process. It is unclear whether the domains identified within a rheumatology context are relevant, comprehensive, or comprehensible when applied in other healthcare contexts, and therefore whether the rheuCOS-SDM is transferable to the assessment of a variety of SDM interventions tailored to and interacting with a range of patient populations, healthcare settings and contexts.

Implementation of SDM interventions across diverse contexts is likely to require tailoring and adaptation of SDM processes through the utilisation of intervention components that interact with the populations and settings in which they are delivered (40). For example, different clinical contexts may require different types of healthcare decisions to be made or bring additional complexity into the patient-practitioner interaction (e.g., for patients who live with multiple long-term conditions). There may be differences in pathways of care and different timeframes for decision-making because of variation in healthcare organisation. There may be differences in societal or cultural expectations, and differences in resource provision because of variation in health care policy. All of these contextual factors influence the implementation of the components of SDM interventions, as well as any assessment of their effectiveness.

#### Aim

The aim of this study is to develop a core outcome set (COS) to evaluate SDM interventions. The COS will be applicable across diverse populations and healthcare contexts, to all types of SDM interventions targeting individuals acting at various levels of a patient-centred healthcare system. The COS will be relevant to patients and caregivers, health care professionals, health services and systems, and wider society. The COS will support consistent outcome selection, reduce reporting heterogeneity, and enable more robust syntheses and economic analyses of SDM interventions. This will enable meaningful health technology appraisal and impact on healthcare policy. We define this new COS as the COS-SDM.

#### Objectives

i. Identify new information sources (to build on the rheuCOS-SDM ‘long list’ of outcomes)
ii. Extract a ‘long’ (extensive) list of outcomes from new information sources, and, where possible, map onto rheuCOS-SDM domains
iii. Gain international consensus on new and existing domains to be included in the COS-SDM
iv. Create a ‘short’ (refined) list of candidate outcome domains
v. Obtain consensus to finalise the COS-SDM

## Methods and Analysis

### Study design

The scope of the COS-SDM (see Box 1) was defined through consultation with a panel of international, multidisciplinary interest holders relevant to a broad range of healthcare contexts (including experts in SDM, health care professionals, public/patient partners). Consultations included two online meetings with structured discussions.

Interest holders include:

- Patients, carers, public members, patient organisations, and patient advocacy groups representing lived experiences of a wide range of healthcare decision-making
- Healthcare and allied health professionals (e.g., doctors, nurses, dentists, dieticians, occupational therapists, paramedics, physiotherapists, psychologists, healthcare educators) involved in the care or decision-making process that includes patients with a wide range of healthcare conditions
- Academic experts in SDM (e.g., trialists, educators, decision scientists, developers of SDM interventions and/or measurement instruments), ethics, law, outcomes methodology, implementation science and health economics.
- Representatives and decision-makers from policy, guideline development, HTA, commissioning bodies or global non-governmental organizations (NGOs)
- Journal editors
- Research funders

#### Box 1

The scope of the Core Outcome Set

**Health conditions**

All health conditions and populations who are involved in healthcare decisionmaking. This can therefore include decisions related to undertaking tests and procedures (e.g., used in screening and diagnosis), making lifestyle choices (e.g., stopping smoking, using physical activity programmes), considering treatment (e.g., medications, surgery, physiotherapy, counselling), or for advanced care planning. The COS will be inclusive of decision-making in maternity care, while recognising that pregnancy may not be considered a health condition, for paediatric care in which adults are the decision-makers, and in situations where there is the need for important others (e.g., carers or relatives) as proxy decision-makers (e.g. where another person participates in SDM on behalf of a patient who is not cognitively able to do so).

**Interventions**

All types of interventions aimed at facilitating SDM in clinical and non-clinical contexts, with components that may target patients/public, health and allied health professionals, patient pathways within health systems, or have a combination of interacting components. Examples of interventions include patient decision aids, decision coaching, question prompt lists, training and feedback programmes for healthcare providers, educational programmes for patients, service changes designed to support patients and/or clinicians during the decision-making process, or any combination of these. (This list is not exhaustive.)

**Context of use**

The COS will be recommended for use in clinical and non-clinical comparative effectiveness research (e.g., randomised controlled trials) and observational, i.e., non-interventional, studies.

**Figure 1.**
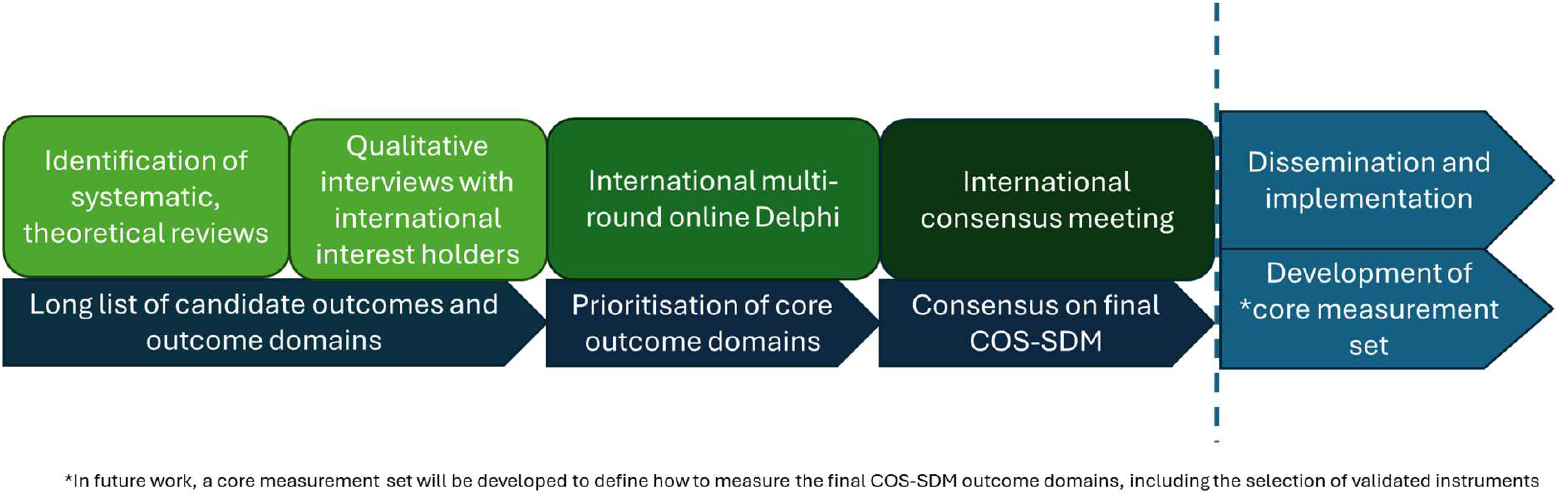
COS-SDM study flow chart.

We will adhere to methods for COS development outlined in the COMET handbook and Core Outcome Set-STAndards for Development (COS-STAD) guidelines (37) and follow guidelines for the reporting of consensus studies (including Delphi) (41). See Figure 1 for a study flow chart. Key steps will include:

- Production of a long list of candidate outcome domains, utilising both evidence synthesis and qualitative research methods (objectives i and ii)
- Prioritisation of core outcome domains utilising a sequential two-round online Delphi (objective iii)
- Reaching consensus on the final outcome set through modified nominal group techniques (objective iv).

This project has been registered in the COMET database (www.comet-initiative.org/Studies/Details/3586).

#### Patient and Public Involvement and Engagement

Patient and public involvement and engagement (PPIE) is central to the development of the COS-SDM. A patient partner (MS) serves as a co-investigator and member of the study delivery team to ensure that patient perspectives remain integral throughout. An international patient/public advisory group (PPAG), comprising patient/public partners from their respective countries has been convened to provide input at various key stages of the project. MS (with support from the study management team) will lead the PPAG activities, monitor progress against milestones, help shape the conduct of consensus activities, and serve as a key link to the executive group by taking part in executive group meetings. The PPAG have already shaped the design of this project. Individual PPAG members have reviewed and co-authored this protocol and all PPAG members are named in the Acknowledgements section. The PPAG will continue to be involved throughout the planning, design, conduct, and dissemination of this study. The PPAG will have significant input and oversight during the development of and recruitment for consensus activities. For example, the group will review outcome domains for relevance and comprehensiveness and assess the Delphi survey for clarity and comprehensibility. Their expertise will be sought when disseminating the questionnaires, supporting recruitment through their international networks and organisations. All patient and public-facing materials will be co-developed with PPAG input. The perspectives of the PPAG will be given equal weighting to the perspectives of the executive group during interpretation of data, to inform decision-making by the project co-leads and the study delivery group. All PPAG activities and the impact that they have on the study will be captured in an Impact Log and reported (42).

### Data collection and analysis

#### Production of a ‘long list’

To produce a ‘long list’ of candidate outcomes and outcome domains we will draw on the data sources used in the development of rheuCOS-SDM along with additional sources that will be used to identify other potential outcomes/domains that were beyond the scope of rheuCOS-SDM. Additional sources will include relevant existing literature as well as primary qualitative interviews. Examples of data sources are presented in table 1.

**Table 1.** Example data sources.

|  |
| --- |
| **Sources used to develop rheuCOS-SDM** |
| Systematic review of SDM domains |
| Cochrane systematic review of outcomes of studies evaluating patient decision aids |
| Interviews with patients/professionals in a rheumatology context |
| Survey of patients/professionals in a rheumatology context |
| **Additional sources** |
| Cochrane review of interventions to increase the use of SDM 2018 |
| Cochrane systematic review of decision coaching for people making healthcare decisions 2021 |
| Scoping review of long-term outcome of SDM on patients, professionals and health services 2024 |
| Theoretical papers describing potential outcomes of SDM process |
| Scoping review of decision aids for health promotion 2021 |
| A framework for conducting economic evaluations when using patient decision aids in health care decision making 2015 |
| Supplementary qualitative, semi-structured interviews with a variety of international interest holders, including, e.g., patients, SDM experts, policymakers, clinicians and allied health professionals |

##### Review of existing literature

There are a high number of existing evidence syntheses related to SDM; we will focus on identifying systematic and scoping reviews. These have to investigate (i) the effects of SDM interventions, or (ii) the impact of implementing SDM and (iii) provide a summary of outcomes from included studies. Review papers also have to have a broad scope so that results can be generalisable to a range of healthcare decisions (i.e., investigate SDM interventions in general, rather than apply to a specific disease or condition only) or focus on one of the broader areas of interest (e.g., decisions related to health promotion/prevention, screening decisions) in line with the scope of this study. Additional data sources will be identified through applying the keyword ‘shared decision making’ to the Cochrane Library database of systematic reviews. Supplementary targeted searches will be conducted in Google Scholar to identify other reviews. Data sources known to the international executive group will be reviewed for eligibility and included if they provide new information (i.e., report additional outcomes not identified through previous data sources).

##### Primary qualitative interviews

Semi-structured, one-to-one interviews will be conducted with a diverse range of relevant international interest holders. Interviews will elicit insights into further outcome domains to be added to the long list as well as perspectives on the wording of these outcome domains, and their descriptions, within the Delphi survey.

A target sample of 20 participants is comparable to studies using similar methodologies and deemed of a size to achieve a breadth of perspective and provide ‘information power’ surrounding the core objectives of this study (43, 44). Sampling will be by geography (i.e., to ensure inclusivity across all continents), experience, expertise, and personal characteristics. A targeted approach will ensure that participant characteristics match those outlined in our list of interest holders. Identifiable participant characteristics will not be reported. Patients and members of the public will be approached through existing patient and public involvement networks linked to members of the research team, by sending an advert via email to individuals who have previously expressed interest in this field of research. Experts in SDM will be approached similarly, by email, using the networks of the International Shared Decision-Making Society (ISDM). Snowballing will then be used to target clinicians and academics linked to country or regional leads for SDM research.

Interviews will be conducted in English, and online, by one researcher (JB – an academic primary care physician and clinical lecturer with methodological expertise that includes qualitative research). Written consent will be obtained and verbally confirmed at the outset of interviews and participants will be given the opportunity to ask questions before providing their consent. Transcription software will be utilised, supplemented by audio recordings to check and correct transcripts. Any video-recorded data will be destroyed with only audio data retained. Transcripts will be pseudo-anonymised before analysis.

A topic guide has been developed to match the objectives of the study, with an introductory question to ask participants about what SDM means to them and any previous experience or expertise with SDM. The topic guide will be piloted and amended as necessary during the first three interviews. A pragmatic constant comparative approach (45) will be employed for the thematic analysis of interview transcripts. Coding will be carried out using NVivo software (version 14.23.1). Two researchers (JB, and TN – a resident doctor with experience in qualitative methods and thematic analysis) will independently code a subset, compare, and if agreement is reached coding will continue by a single researcher. Coding will be both deductive and inductive. Deductive coding will follow the broad themes contained within the semi-structured topic guide, to directly influence the long list of outcome domains. Inductive coding will capture new themes and theory not previously considered by the research team. As the interviews aim to gather additional insights and outcomes from diverse international stakeholders to build on and validate the existing list of outcome domains, data saturation is not the goal. Saturation is unlikely to be achieved given the international breadth of perspectives (46). Individual participants will not be invited to provide feedback on results. However, members of the PPAG, along with the multidisciplinary, international executive group, will provide a perspective to guide interpretation and presentation of findings.

##### Data extraction

Outcomes from all data sources will be extracted verbatim into a long list and mapped, where possible, onto domains identified in the rheuCOS-SDM. Outcomes that do not align with existing domains will be reviewed and grouped into further outcome domains (guided by the executive steering group and the PPAG) to be presented separately during the consensus process. The process of grouping outcomes will be guided by existing frameworks, including the OMERACT filter 2.1 (47) and the COMET outcome taxonomy (48), to create a short list of candidate outcome domains.

The executive group, and the PPAG, will review the short list of all candidate outcome domains for comprehensiveness, comprehensibility, and relevance to the new context. Revisions will be made, based on their feedback.

#### Prioritisation of core domains

All outcome domains will be operationalised into an online Delphi questionnaire. Draft surveys will be piloted and examined for face validity by members of the PPAG. The questionnaires will be hosted online using appropriate software (e.g., REDcap). Questionnaires will be developed in English. The executive group considered multi-lingual COS development; however, a single language option was preferred to retain conceptual consistency during the survey development, and because there were concerns about feasibility within the resources available for the study.

A two-round Delphi process will be undertaken inclusive of all interest holders relevant to this project. Following responses, participant data will be separated into at least two Delphi panels depending on which interest holder group they represent. As a minimum, participants may be divided into patients, carers and family members (panel 1) and professionals including clinicians and SDM researchers (panel 2), to be able to assess differences in priorities and to avoid overrepresentation of a single group.

Participants will be invited to complete two consecutive rounds of the survey. In Round 1, participants are asked to provide sociodemographic details to determine the relevant panel group. During both rounds, participants will be asked to rate the importance of assessing each outcome domain using a Likert scale based on the Grading of Recommendations, Assessment, Development and Evaluations (GRADE) scale (49). This means scoring will be undertaken using a 9-point Likert scale with 1-3 labelled ‘not essential’, 4-6 labelled ‘important but not essential’ and 7-9 labelled ‘absolutely essential’ (49). There will also be an “Unable to answer” option for each question with the ability for the participant to enter free text to explain why they were not able to score the item. A single ‘free text’ response option at the end of the survey will be included to allow participants to suggest any additional domains. Participants will be informed that they should not base their responses to the survey on whether or not a validated measure is already available for a particular outcome domain. Outcome domains will be worded neutrally, in a way that can be interpreted bidirectionally without influence on the participant, i.e., ‘patient trust’ as opposed to ‘more patient trust’ or ‘less patient trust’.

New potential outcome domains, identified by at least 10% of respondents through the free text response, will be included in subsequent Delphi rounds. Only participants who complete Round 1 will be invited to Round 2. All questions will be mandatory to minimise missing data. Questions presented in Round 2 will be accompanied by feedback from Round 1 responses and participants will be asked to reconsider their response in light of this feedback. Feedback will consist of the average score for each question for each participant panel (i.e., median score per outcome domain/question) and the individual participant’s own previous rating. All items will be retained in full between survey rounds 1 and 2. All items still retained after Round 2 of the Delphi survey will be taken forward to the consensus meeting where the final COS will be agreed (37).

Participants will be recruited through adverts disseminated by established international SDM societies and OMERACT networks. Those approached will not be offered any financial incentive to participate. Participants will be purposively approached to ensure variation in key characteristics including sex, geographical location, ethnicity, age, healthcare experience or professional/clinical expertise (depending on the interest holder group). Participant characteristics will be reported using the PROGRESS+ framework (50). Round 1 Delphi survey participants will be asked to confidentially provide their email address which will be used to facilitate recruitment for Round 2 Delphi survey. A direct email will be sent which will include an embedded link to the second round. A minimum of 150 participants per panel will be sought, in line with previous COS development studies (37). There will be no maximum sample size, and a weighted percentage will be considered should participant numbers be uneven across the different panels (e.g. patient/carer vs. professionals) (37).

#### Reaching consensus

##### Analysis of survey data

Participant response scores for the Delphi process will be analysed using descriptive statistics, adhering to the GRADE criteria. Analyses will be conducted separately for each panel and average scores calculated (i.e., the median score of each panel per outcome domain/question). Results for each panel will be presented visually as feedback in Delphi Round 2. Final Delphi survey scores will again be subject to descriptive analyses with any changes in participant scores between rounds examined.

Response statistics and participant characteristics will be examined using descriptive statistics and presented in tabulated format by panel group. Attrition between Delphi survey rounds will be examined for potential bias in the different participant panels, comparing scores between participants who have completed only one Delphi survey round versus participants who have completed both Delphi survey rounds.

All analyses will be conducted with the help of a statistical software package (e.g., Stata).

##### Defining consensus

Recommended definitions of consensus will be followed during Delphi Round 1 (see Box 2) (37). Definitions may be revised for Round 2 analyses, in case stricter criteria (e.g., higher percentage thresholds) are necessary because of a high number of participants rating outcome domains as ‘absolutely essential’ [7-9]. Any amendments to consensus definitions will be discussed with the executive group and PPAG.

###### Box 2

Consensus criteria

- Consensus IN is defined as >70% of the total participants rating the outcome domain as 7-9 (‘absolutely essential’) and <15% rating the outcome domain as 1-3 (‘not essential’). This means the outcome domain will be taken forward to the consensus process.
- Consensus OUT is defined as >70% of participants rating the outcome domain as 1-3 (‘not essentia|l’) and <15% rating the outcome domain as 7-9 (‘absolutely essential’). This means the outcome domain will not be taken forward to the consensus meeting.
- All other combinations of response results are defined as NO CONSENSUS

##### Modified nominal group techniques

Multiple international online consensus meetings will be conducted independently to discuss the results of the Delphi Round 2 results. The number of meetings will be determined in discussion with the executive group, considering the geographic representations and involvement of diverse interest holders. For example, it is anticipated that three meetings will be held to accommodate time zones in North and Latin America, Europe and Africa, Australia and Asia.

Recruitment for the consensus meetings will be targeted and facilitated through international SDM societies and OMERACT networks, and key informant approaches. It is anticipated that no more than 25 participants, with fair representation of the interest holder groups list above, will participate in each consensus meeting. A secure video conference platform will be used to host the meetings.

At each meeting, results will be presented by an independent chairperson with an understanding of SDM interventions. The Chair will lead structured discussions about the proposed inclusion/exclusion of domains from the COS, based on the consensus definitions. This will include discussions about combining outcomes depending on similarity. Any divergence in consensus between the two panel groups will be discussed. Interactive, anonymous voting will be used to gain consensus on proposed domains that achieve ‘NO’ consensus from the Delphi rounds. Those domains achieving ‘IN’ or ‘OUT’ status in the Delphi will simply be presented as such, and voting on these will only take place if there is sufficient objection to the status of these domains by the group.

The results of the multiple online consensus meetings will be synthesised, reported, and considered at a hybrid (both online and in-person) symposium at an international conference (e.g. the ISDM 2026 conference, Dartmouth, New Hampshire, USA) attended by patient and professional interest holders from diverse backgrounds. This process will result in the reporting of the final COS-SDM.

## Data Availability

All data produced in the present work are contained in the manuscript

https://www.comet-initiative.org/Studies/Details/3586

## Ethics and Dissemination

### Ethics

Research ethics approval has been granted in the UK (University of Bristol Faculty Ethics Committee ref: 7741; University of Exeter Health and Community Sciences Ethics Committee ref: 8207624). Written informed consent will be obtained digitally from all participants in the Delphi and the consensus meetings.

### Dissemination

The final COS will be disseminated by presentation at international SDM conferences and results of this study will be published in a peer-reviewed journal, facilitated through a partnership with BMJ EBM. Tailored to the needs of a wide, diverse audience, the delivery group will consider the production of Plain Language summaries, infographics, and/or translated materials, as appropriate. Further dissemination is planned through the patient/public advisory group, ISDM, OMERACT, and executive group channels, to publicize the COS to patient groups, funders, journal editors, regulatory bodies and HTA boards internationally.

## Author contribution

### Co-leads

Toupin-April, Karine; McNair, Angus

### Project delivery

Hoffmann, Christin; Butterworth, Joanne; Naye, Florian; Smith, Maureen; Nyamapfene, Taona; Lawday, Samuel

### Executive team

Avery, Kerry; Bekker, Hilary; Bravo, Paulina; Décary, Simon; Edwards, Adrian; Elwyn, Glyn; Engelhardt, Ellen; Franco, Juan; Garvelink, Mirjam; Giguère, Anik; Härter, Martin; Hoffmann, Tammy; Kienlin, Simone; McCaffery, Kirsten; Noordman, Janneke; Olling, Karina; Perestelo-Perez, Lilisbeth; Pieterse, Arwen; Scheibler, Fülöp; Sepucha, Karen; Stacey, Dawn; Ubbink, Dirk; Valentine, Kathrene; Volk, Bob; Wehking, Felix; Yoo, Sang-Ho.

### Authors who are also members of our Patient and Public Advisory Group

Bulbeck, Helen; de Wit, Maarten; Martí, Norma; Snede, Lisbeth

### Members of the wider COS-SDM study group

Cole, Amy; Finderup, Jeanette; Geary, Katie; Gunn, Christine; Hou, Wen-Hsuan; Housten, Ashley; Kim, Min Ji; Pacheco-Brousseau, Lissa; Tian, Cindy Yue,

## Acknowledgements

**Patient and public advisory group**

Andersen, Vibeke; Bulbeck, Helen; Chen, Xuanna; de Wit, Maarten; Ebai-Atuh, Ndi Euphrasia; Martí, Norma; Snede, Lisbeth; Stensdal, Lilli-Ann

